# Understanding the prevalence of psychiatric disorders in people with cerebral palsy: A systematic review protocol

**DOI:** 10.1101/2021.03.23.21254205

**Authors:** R. Eres, I. Dutia, M. Walsh, M. Mulraney, S.M Sawyer, D.S Reddihough, D. Coghill

## Abstract

**Background:** There is an abundance of research investigating physical health outcomes in people with cerebral palsy, however evidence from studies investigating psychiatric disorders in this group is less well developed. Specifically, there is little research synthesising the prevalence rates of psychiatric disorders in people with cerebral palsy, and there is no summary of the instruments used to measure this. This systematic review will summarise the current body of evidence investigating psychiatric disorders in people with cerebral palsy, as well as the measurement or diagnostic instruments used to measure psychiatric disorder.

**Findings:** To be included in the review, articles must report on psychiatric disorders in people with cerebral palsy or use standardised tools that allow for clinical thresholds to be derived, written in English, and published after 1980. A comprehensive, electronic database search strategy will be implemented targeting relevant papers indexed in MEDLINE, PsycINFO, Cochrane Library, and PubMed. Searches will include terms that relate to psychiatric disorders (E.g., mood disorders, anxiety disorders, ADHD, autism) in people with cerebral palsy. Data will be extracted based on age, psychiatric disorder, and cerebral palsy severity. We will summarise the findings as a narrative and will provide overall prevalence estimates for psychiatric disorders.

**Discussion:** The systematic review will provide novel insights into the prevalence of psychiatric disorders in people with cerebral palsy. It will also provide recommendations about the types of instruments that have been used and implemented in health research related to cerebral palsy. It will guide future work on interventions and primary research in clinical sciences and allied health disciplines.

## Background

Cerebral palsy (CP) is a group of permanent disorders of the development of movement and posture, causing activity limitations that are attributed to nonprogressive disturbances that occur in the developing foetal or infant brain (Bax et al., 2005). The location and severity of these nonprogressive disturbances has implications for the functionality and quality of life that people with CP experience. Some people with CP may solely experience a minor motor disruption but are otherwise completely independent, while others may have severe disabilities that impact every area of their functioning. Thus, CP can be considered a spectrum of functional capacity that varies between each person diagnosed with this disease.

Given the heterogenous nature of CP presentations, there has been a substantial amount of research investigating the physical health concerns of people with CP. Much of this research is in children with far fewer studies investigating outcomes in adult populations. Regardless the general consensus stands that people with CP have worse physical health than the general population. For example, people with CP are at increased risk of developing diabetes, heart disease, and stroke (Peterson et al., 2015), and have higher rates of hospital admissions and access acute inpatient services for emergency health care more than people without CP (Morgan et al., 2020). People with CP have an increased risk of early mortality due to ischaemic heart disease, cerebrovascular disease, respiratory diseases, and circulatory disorders (Ryan et al., 2019). They also experience more fatigue, insomnia, and pain compared with their able-bodied peers (Vogtle et al., 2009; Van der Slot et al., 2012).

While there is a significant body of work investigating the physical health of people with CP, mental health outcomes are less well studied. When mental health is studied in CP it has tended to be at the symptom level. There is for example evidence to suggest that people with CP report higher levels of anxiety and depression symptoms compared with their able-bodied peers (Jarl et al., 2019; van Gorp et al, 2019). However, what is less known is the prevalence of common psychiatric disorders in this population across the lifespan. Recently, there have been some investigations that show an increase in prevalence rates for mood, anxiety, and psychotic disorders for people with CP compared with the general population (McMorris et al., 2021; Whitney et al., 2019), but a comprehensive examination of these prevalence studies remains to be completed. Further, there is currently no investigation that summarises the measurement instruments used to assess psychiatric disorders or mental health symptomatology in people with CP. This is important as while many people with CP are able to access the same standard instruments that are typically used in general population and clinical studies others, particularly those with communication difficulties and intellectual disabilities are likely to find this difficult.

Thus, the current protocol describes the processes for conducting a systematic review investigating the prevalence rates of psychiatric disorders in people with CP across the lifespan, as well as the measurement instruments used to assess psychiatric disorders in this population. This systematic review holds importance for understanding the mental health status of people with CP and will help support the development of assessment tools that can accurately and effectively assess mental health disorders in people with CP.

## Methods and Design

This secondary data synthesis will be a systematic review of studies reporting psychiatric disorders present in people with CP. We will follow the transparent reporting of systematic reviews and meta-analysis (PRISMA) guidelines to conduct this systematic review. The primary outcomes of this systematic review will include any psychiatric diagnosis (e.g., mood disorders, anxiety disorders, autism spectrum disorders, attention deficit hyperactivity disorder, psychotic disorders) from any known diagnostic method (e.g., ICD classification, DSM classification, semi-structured clinical interviews, including SCID, MINI, and K-SADS-PL). Studies using standardised clinical cut-off scores to determine clinical diagnosis will also be accepted. Additional demographic characteristics will be collected to determine whether there are changes in prevalence between age groups across the lifespan.

In the sections that follow we will describe I) inclusion and exclusion criteria, II) literature search protocols, III) study screening and selection processes, IV) data extraction, V) assessing risk of bias procedures, and VI) analysis of findings.

### I. Inclusion and exclusion criteria

Studies will be included in the systematic review if they meet the following criteria: I) report on psychiatric disorders in people of any age with CP either through primary or secondary research, II) reports on mental health symptomatology using a measurement tool that has validated clinical cut off scores (e.g., Centre for Epidemiological Studies – Depression with a standardised cut-off score of 16 to indicate depression), III) is written in English, and IV) was published between 1980 and 2021.

Studies will be excluded from the systematic review if: I) non-human animals are recruited, II) studies that include participants with CP but where those participants’ data are indistinguishable from other child-onset developmental disorders (e.g., neurological disorders more broadly, or congenital spinal malformations which may include spina bifida, CP, and muscular dystrophy), III) studies published prior to 1980, and IV) studies published in a language other than English.

### II. Literature search protocols

We will use two search strategies to identify literature suitable for this review article. First, we will complete an extensive electronic bibliographic database search. Second, we will conduct a backward literature search using the reference lists of previously published synthesis articles.

#### II.I Bibliographic database search

Literature searches will be conducted in several databases including PubMed, PsycINFO, Cochrane Library, and MEDLINE. Search terms will be tailored to the specific database that we are examining at the time, and we will receive support from a librarian to assist with specific search strategies. Initial strategies will include terms that relate to specific psychiatric diagnoses (e.g., depression, anxiety, psychosis) and qualified with Boolean operators to ensure the necessary articles are captured. We will use search filters to ensure that articles retrieved meet the inclusion and exclusion criteria related to date of publication, language, and human participants. We will also use truncations (e.g., * in depress to obtain depression, depressed), wildcards (e.g., ? to accommodate for difference in spelling between British and American spelling), and proximity operators (e.g., adding Adjn to your search “animal Adjn therapy” will provide results of “animal assisted therapy, animal-based therapy, animal-based care therapy).

#### II.II Backward literature search

We will also use a backward literature search strategy to identify relevant articles from studies that have previously been published that may not appear in the databases because they are either in press or very recently published. This will involve reviewing the reference list of published articles and reviewing articles that may be related to the current systematic review.

### III. Study screening and selection process

On completion of the initial search strategy, title review and abstract review will occur independently by two authors. The inclusion and exclusion criteria will be used to categorise studies as either relevant or irrelevant for the review. The two coders, authors RE and ID, will independently screen records for inclusion into the systematic review using covidence software with citations exported to Endnote libraries for the citation record keeping. Coders will be blind to each other’s coding. Disagreements on coding will be discussed between the two coders where they will re-evaluate the inclusion and exclusion of those articles. If no solution is converged on, a third coder (MM) will be included for discussion. This same process will occur for full-text review after the completion of the title and abstract review processes.

### IV. Data extraction

A standardised and pre-piloted data extraction template will be used to extract data from screened studies. Extracted information will include, number of participants included in study, demographic details of CP sample (e.g., gross motor classification system, manual ability classification system, communication function classification system, eating and drinking ability classification system, intellectual disability) and comparator sample if provided, prevalence estimates of psychiatric disorders, and diagnostic tool used to assess psychiatric disorders. Two authors will independently use this template to extract information and will later meet to discuss any discrepancies. A third author will be included if no agreement is reached between the two reviewers. Missing data will be requested from study authors through email correspondence.

### V. Assessing risk of bias

Two authors will independently assess the risk of bias of each article included in the review based on the 10 criteria extracted from an adapted version of the Grades and Recommendation, Assessment, Development and Evaluation (GRADE) tool and Cochrane approaches (Hoy et al., 2012). This information will be collected using a standardised checklist.

The risk of bias tool will specifically assess the external and internal validity of each study included. These are highlighted below.

External validity is assessed against four criteria including:

1. Was the study’s target population a close representation of the national population in relation to relevant variables?
2. Was the sampling frame a true or close representation of the target population?
3. Was some form of random selection used to select the sample, OR, was a census undertaken?
4. Was the likelihood of non-response bias minimal?

Internal validity is assessed against six criteria including:

1. Were data collected directly from the subjects?
2. Was an acceptable case definition used in the study?
3. Was the study instrument that measured the parameter of interest shown to have reliability and validity (if necessary)?
4. Was the same mode of data collection used for all subjects?
5. Was the length of the shortest prevalence period for the parameter of interest appropriate?
6. Were the numerator and denominator for the parameter of interest appropriate?

Disagreements between the assessors will be discussed in full to resolve the discrepancies between assessors. If the two assessors cannot converge on an agreed risk of bias score for a particular paper, a third assessor will be consulted to determine the appropriate risk. The level of risk associated with each article will be reported in the final paper or provided upon request. Risk of bias scores will be presented in a tabulated table with criterion scores presented for each study.

### VI. Analysis of findings

We will provide a narrative synthesis of the extracted data. We will structure the findings in terms of age cohorts that make sense based on the available data. Ideally, we would present the prevalence rates of data across children (6-11years of age), adolescents (12-17 years), young adults (18 – 25 years), adults (26 – 64 years), and older adults (65+ years). If data is available, comparison groups of non-cerebral palsied participants will be included and contrasted with the CP participants in each study that they are included. Potential mediating and moderating factors will be examined as necessary.

## Discussion

This systematic review is a necessary body of work that will provide a detailed summary of psychiatric disorder prevalence rates in people with CP. It will help provide guidance in the establishment of standardised psychiatric assessment tools for this population and will illustrate novel insights into the mental health status of people with CP.

## Data Availability

There is no data available for this systematic review protocol.

